# Evaluation of a prioritization system for mild traumatic brain injury case ascertainment from emergency department records

**DOI:** 10.1101/2024.11.09.24317035

**Authors:** Katherine MacMillan, Tasha Klotz, Noah D. Silverberg

**Affiliations:** Department of Psychology, University of British Columbia, Vancouver, BC, Canada; Rehabilitation Research Program, Centre for Aging SMART, Vancouver Coastal Health, Research Institute; Djavad Mowafaghian Centre for Brain Health

**Keywords:** chart review, electronic health record, electronic medical record, mTBI, mild traumatic brain injury, algorithm, key term, concussion, chart review system, tiered priority screening system, recruitment, mTBI, TBI, emergency department recruitment, chief complaint, reason for visit

## Abstract

**Objective:** To evaluate modifications to a tiered priority chart review system designed to efficiently identify patients with mild traumatic brain injury (mTBI), via medical chart review, that presented to an emergency department or urgent and primary care center (ED/UPCC) for research purposes.

**Methods:** We initially created a tiered priority chart review system and applied it to screening N=17,072 electronic ED/UPCC medical charts in study 1 (Clinicaltrials.gov ID# NCT04704037). Chief complaints with high positive predictive value (PPV) for correctly identifying possible/probable mTBI cases were moved to a higher tier and those with low positive predictive value were downgraded to create a tiered priority chart review system. This revised system was then used in a second research study (Clinicaltrials.gov ID# NCT05365776), and PPV values were calculated for the new sample (N=4,434). The original and revised tiered priority system were compared with respect to overall efficiency.

**Results:** PPV for specific chief complaint key terms varied markedly from 0% to 61% and resulted in an empirically-driven resorting of the priority tiers. After excluding clearly ineligible participants, 49% of charts reviewed in the first study and 60% of charts reviewed in the second study were identified as a possible or probable mTBI. This indicates an improvement in overall efficiency (12%; χ^2^(1)=114.7, p<.001) compared to the original system.

**Conclusion:** The revised tiered priority chart review system was more efficient at identifying patients with mTBI for the purposes of mTBI study recruitment.

## Introduction

Identifying potential participants with mild traumatic brain injury (mTBI) from emergency departments and urgent and primary care centers (EDs/UPCCs) can be an efficient recruitment method for mTBI research studies^1,2,3^. Challenges with using diagnostic codes for this purpose have been well-documented^3,4,5^. An alternative may be to use chief complaint/reason for visit fields. ED/UPCCs have very high patient throughput, making it resource intensive, if not infeasible, to review every chart. Narrowing the selection of charts to review with chief complaints associated with mTBI diagnosis could make this process more efficient. However, it is unclear which chief complaints/reason for visits are most associated with mTBI diagnosis. We describe here procedures used to evaluate, modify, and re-evaluate a tiered priority chart review system. The first iteration of the tiered priority chart review system was applied in a research study that recruited adults with mTBI from EDs/UPCCs (Clinicaltrials.gov ID# NCT04704037). We analyzed the positive predictive value (PPV) of chief complaints/reasons for visit (likelihood of mTBI diagnosis, given the presences of a chief complaint), and used these findings to calibrate the tiered priority system, creating a revised version, by moving those with high PPV to a higher tier and downgrading chief complaints with worse PPV to a lower tier. We then applied the revised tiered priority chart review system to a second study, with a non-overlapping sample (Clinicaltrials.gov ID# NCT05365776). This study recruited patients from outpatient clinics and EDs but only the latter recruitment pathway was analyzed here. We hypothesized that the revised tiered priority chart review system would outperform the original system with respect to overall efficiency (proportion of reviewed charts that resulted in a possible or probable mTBI determination according to the case ascertainment algorithm (Figure 1)).

**Figure 1.**
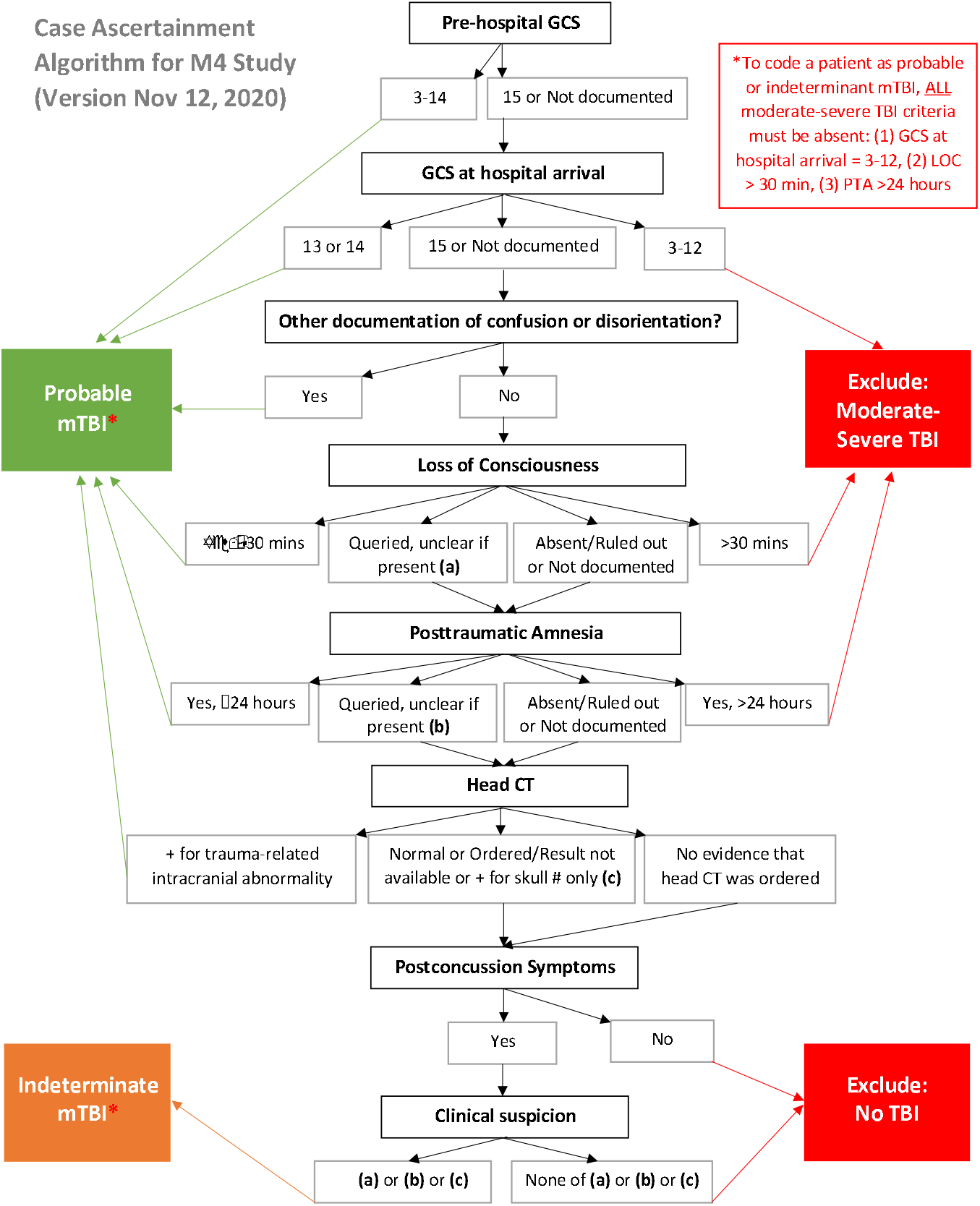
Case ascertainment algorithm. Used in study 1 and study 2 to code emergency department medical charts as probable mTBI, possible/indeterminate mTBI, or Exclude: No TBI or moderate/severe TBI.

## Methods

The original tiered priority system (Appendix A) was created by the principal investigator (NDS) with input from co-authors who had a background in emergency medicine and neurology, for the purpose of identifying potential participants with mTBI for a clinical trial^6^.There were four priority tiers. The highest tier included terms thought to be most associated with possible/probable mTBI diagnosis. The lowest tier included terms that might, in rare cases, be associated with possible/probable mTBI diagnosis, and were expected to have the lowest yield rate. Research assistants were directed to start each shift by reviewing charts with the terms in the highest tier, and as time allowed, move on to lower tier terms. They reviewed charts from the previous 7 days and any chart that had a term present was opened for review.

Research assistants only reviewed the data in the record that was required to fully answer the questions of the case ascertainment algorithm (Figure 1). This included emergency health service (ambulance crew) report, ED/UPCC triaging and assessment notes, diagnostic codes, and radiologist reports when available. Data for both studies was collected in REDCap (Research Electronic Data Capture system).

### Participants

For study 1, electronic charts were accessed from the following platforms: Primary Care/Community Information Systems (PCIS) (Vancouver General Hospital and Richmond Hospital), Clinical & Systems information (CST) Cerner (Lions Gate Hospital, St. Paul’s Hospital, and Mount Saint Joseph’s Hospital), and Intrahealth Profile (City Center Urgent Primacy Care Center and North Vancouver Urgent Primary Care Center). In study 1, participants physical charts were also reviewed at Vancouver General Hospital, Richmond Hospital, and University of British Columbia Hospital. CST Cerner and Intrahealth Profile both used a free text field for chief complaint/reason for visit. For study 1, 17,072 charts, with the date of visit to an ED/UPCC between February 9, 2021 to January 27, 2022, were reviewed using the original tiered priority system.

In the study 2, electronic medical charts were reviewed from: Richmond Hospital, Vancouver Hospital, St. Paul’s Hospital, Lions Gate Hospital, and Mount Saint Joseph’s Hospital using the following platforms: PCIS (Richmond Hospital), CST Cerner (Vancouver General Hospital, Lions Gate Hospital, St. Paul’s Hospital, and Mount Saint Joseph’s Hospital). Each chart that was reviewed included a chief complaint or reason for visit that was recorded by the site staff who did the intake for the patient. There were 4,434 charts that were reviewed with their date of hospital visit between November 18, 2022, to August 27, 2024

### Case ascertainment algorithm

In study 1 and study 2, charts that did not pass a “quick screen” did not have the case ascertainment algorithm (Figure 1) applied to the remainder of their chart and no determination of mTBI was made. These criteria were: (1) home address outside of BC (2) no plausible mechanism for mTBI (3) pre-existing or unstable neurological condition or severe mental illness present (4) outside of the age criteria (18-69 years old). For study 2 this also meant no record was created in REDCap (Research Electronic Data Capture system).

A probable mTBI assignment was given to charts that had one or more of the criteria for mTBI based on the World Health Organization (WHO) guidelines for the diagnosis of a concussion by medical professionals: (1) an initial GCS score of 15 (or undocumented) or a 13-14 when presenting at the hospital, (2) documentation of confusion or disorientation after the injury (3) a witnessed or self-reported loss of consciousness for 30 minutes of less (4) post traumatic amnesia that lasted less than 24 hours (5) a positive CT scan for an intracranial injury.

A possible/indeterminate mTBI assignment was given to a chart that had one or more of: (1) a loss of consciousness (LOC) that was “queried, unclear if present”, (2) post-traumatic amnesia (PTA) that was “queried, unclear if present”, (3) a normal CT scan, paired with two or more post-concussion symptoms (ex. headache, nausea/vomiting, dizziness, balance problems, fatigue, memory problems, concentration problems, or sensitivity to light or noise, etc.). A possible/indeterminate mTBI was also assigned if an ED/UPCC physician provided a mTBI diagnosis on the chart, regardless of the case ascertainment algorithm outcome.

Moderate/severe TBI assignments were given to charts that (1) had a GCS at the hospital between 3-12, (6) had a LOC greater than 30 minutes, (7) had PTA greater than 24 hours.

Chart were excluded for not having a TBI if they did not meet any of the previously mentioned criteria for probable, possible/indeterminate, or moderate/sever TBI and (1) LOC was absent or ruled out (2) PTA was absent or ruled out (3) No evidence for a head CT (4) No post concussion symptoms or (5) no clinical suspicion present.

### Key term coding

As many of the chief complaint/reasons for visit collected were written in free form from ED/UPCC staff, a consistent term needed to be identified for each chief complaints/reasons for visit before the data could be analyzed. To do this every chief complaint/reason for visit was manually reviewed and was recoded with a “key term” to replace the words captured for their chief complaint/reason for visit.

During the re-coding process, several assumptions were made – LOC was assumed to stand for loss of consciousness, HI was assumed to stand for head injury, MVC was assumed to stand for motor vehicle collision, and MVA was assumed to stand for motor vehicle accident. Nausea and vomiting were combined into the key term “Nausea/Vomiting” because they appeared together so often. In reference to bike accidents, mountain bikes and bicycles were counted as separate key terms. Additionally, head, face, and neck terms were all kept separate and not combined. For our definition, when the term head was mentioned in a chief complaint, we assumed it was referring to the skull area or the forehead if not already said simply as “head” in the chief complaint. Additionally, in review, there were many charts that appeared as “Minor head injury” – this was coded as the key term “Minor head injury” rather than “Head injury,” because it appeared by itself so often.

Key terms were assigned based on the chief complaint/reason for visit recorded for each participant. For example, if a chief complaint/reason for visit mentioned that the patient had sustained a concussion, the key term would be “Concussion”. Some chief complaints/reason for visit included more than one term/complaint/reason that could be used as a key term, and thus a priority list was created to decide which key term should be used based on level of importance (Figure 2). More clinically serious condition would be listed as the main key term over less serious condition. For example, if a chief complaint mentioned both concussion and vomiting, the key term would be concussion. Figure 2 demonstrates 1 being the highest priority key term, and larger numbers reflecting lower importance.

**Figure 2.**
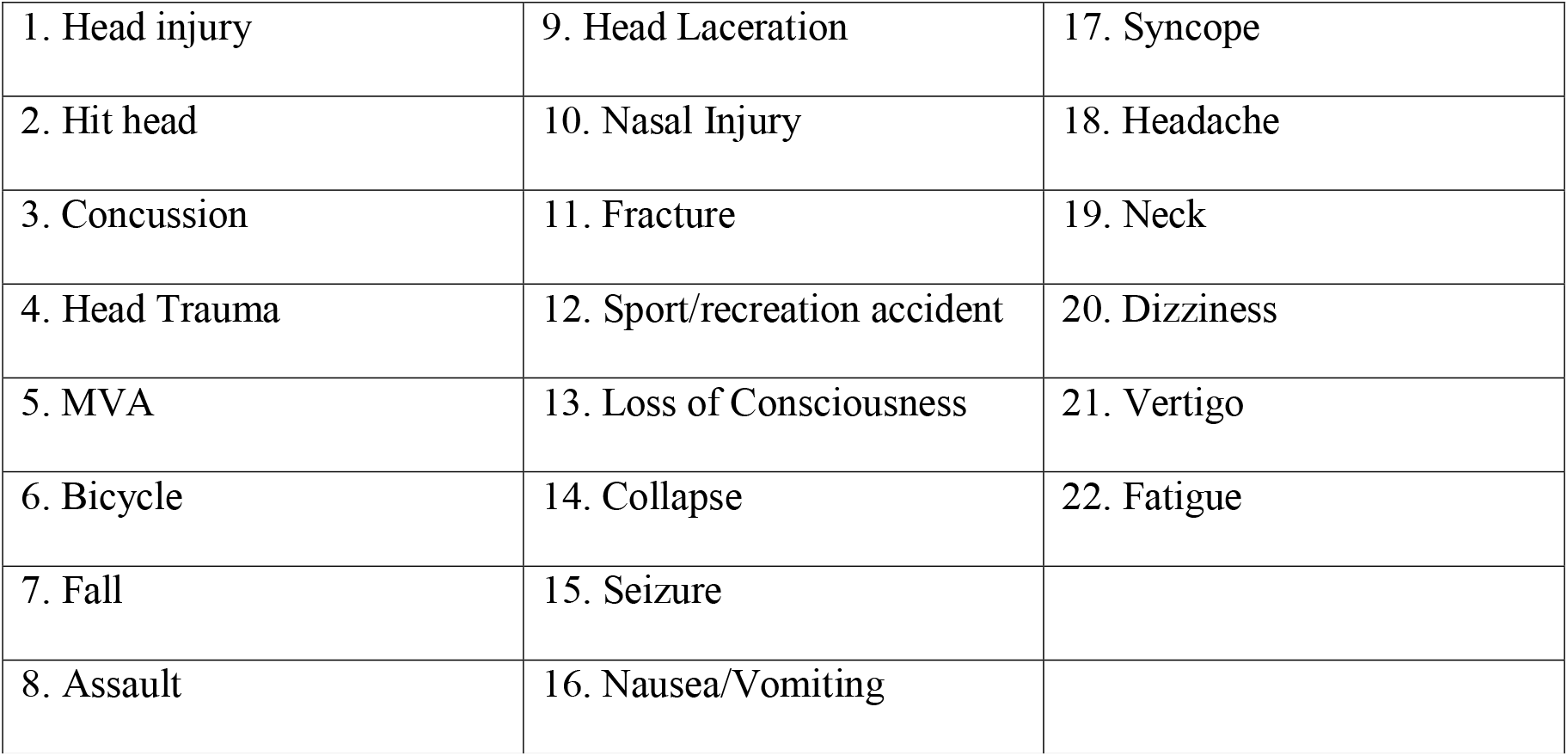
Hierarchical list. Used for deciding which most common terms in a chief complaint should be the key term. This is to be used when multiple potential key terms are present, to decide which term in the chief complaint should be the key term.

### Statistical analysis

From study 1, 16,997 cases were used during analysis. After recoding all chief complaints/reasons for visit with a key term, calculations were completed on all key terms identified. We calculated the Positive Predictive Value (PPV), Sensitivity (Se), Specificity, and Negative Predictive Value (NPV) for each key term. The calculated positive predictive value indicated the probability of a patient having a possible or probable mTBI based on the case ascertainment algorithm plus the presence of a key term (which was based on their chief complaint/reason for visit captured during chart review). A full example using real data for the key term “Head injury” is shown in Figure 3.

**Figure 3.**
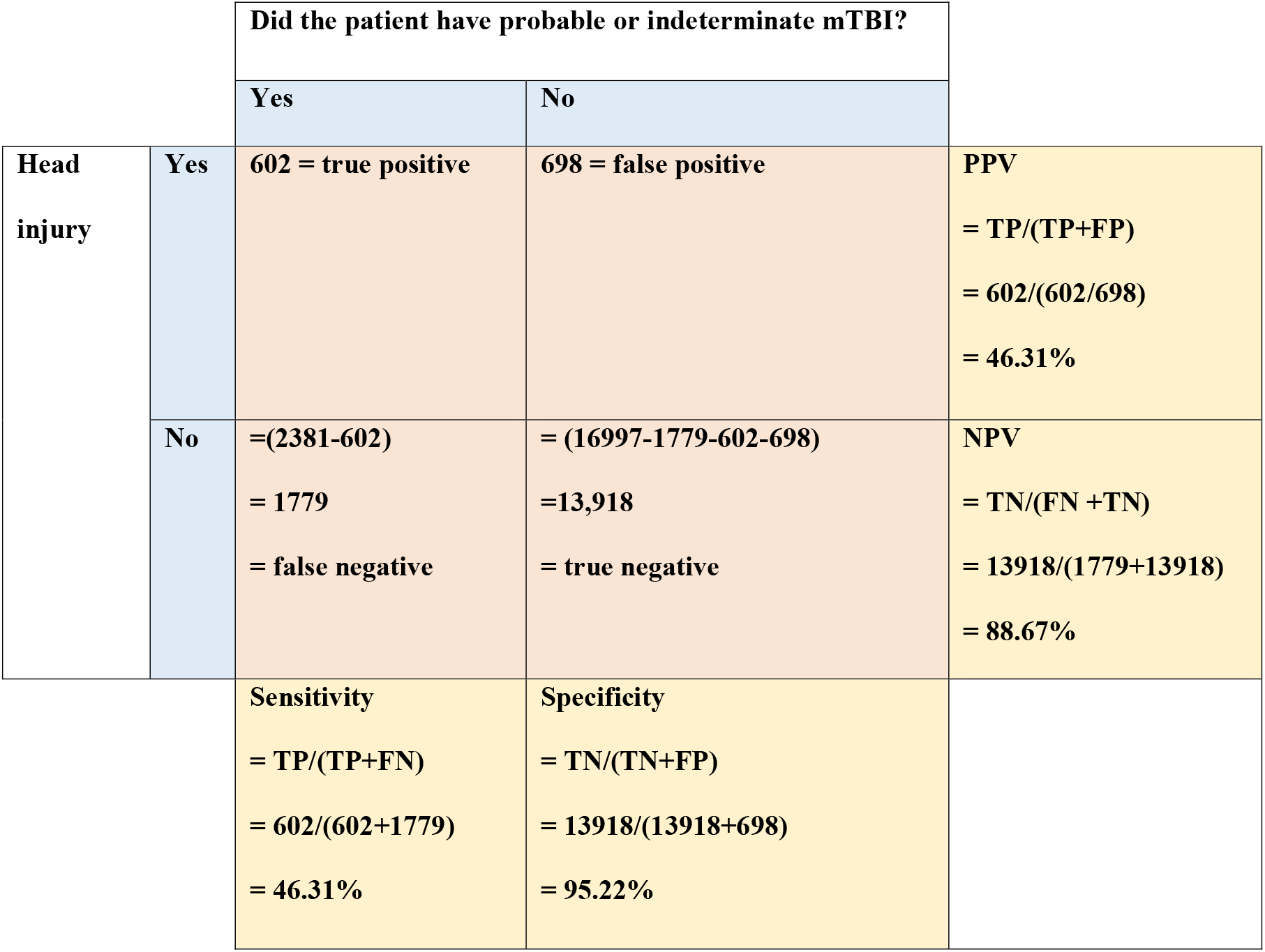
Head injury calculation. Example calculations of the PPV, NPV, sensitivity, and specificity of the key term “head injury”. 1,300 charts out of 16,997 had the key term of head injury in their chief complaint. 602 of those people sustained a possible or probable mTBI based on chart review (the true positive value). 698 of charts that had head injury in their key term were determined to be excluded for the study during chart review (the false positive value). 2,381 people total sustained a possible or probable mTBI, and thus the false negative number 1,779 was obtained by taking the total number of people and subtracting the number of people who had head injury for their key term and sustained a probable or indeterminate mTBI, (2,381-602=1,779). The true negative value was obtained by taking the total number of charts reviewed, and subtracting TP, FP, and FN.

### Creating a New Tiered Priority Chart Screening System

To modify the tiered chart review priority system used in study 1, we considered PPV values for each key term relative to others. Key terms of charts were sorted by their PPV values into five tiers to begin the organization of the new tiered chart and narrow down which key terms would go where in the new system. The categories were: PPV above or equal to 20% (Tier 1), PPV above or equal to 11% up to 20% (Tier 2), PPV above or equal to 4%, up to 11% (Tier 3), PPV above 0%, up to 4% (Tier 4), and PPV = 0% (Tier 5). PPV for all key terms are displayed in Figure 4, sorted by PPV. In the final version of the revised chart review priority system (Figure 5), we had a total of three priority tiers. The first priority tier included the PPV range of above or equal to 20%, the second tier had PPV above or equal to 4% up to 20%, and the third was PPV above 0% up to 4%. In this version, the notation “*” was used to indicate additional common descriptors (or specifiers) of the key term. While these additional common descriptors were not included in our calculations, they were included to assist the research assistants in their determinations while doing chart review.

**Figure 4.**
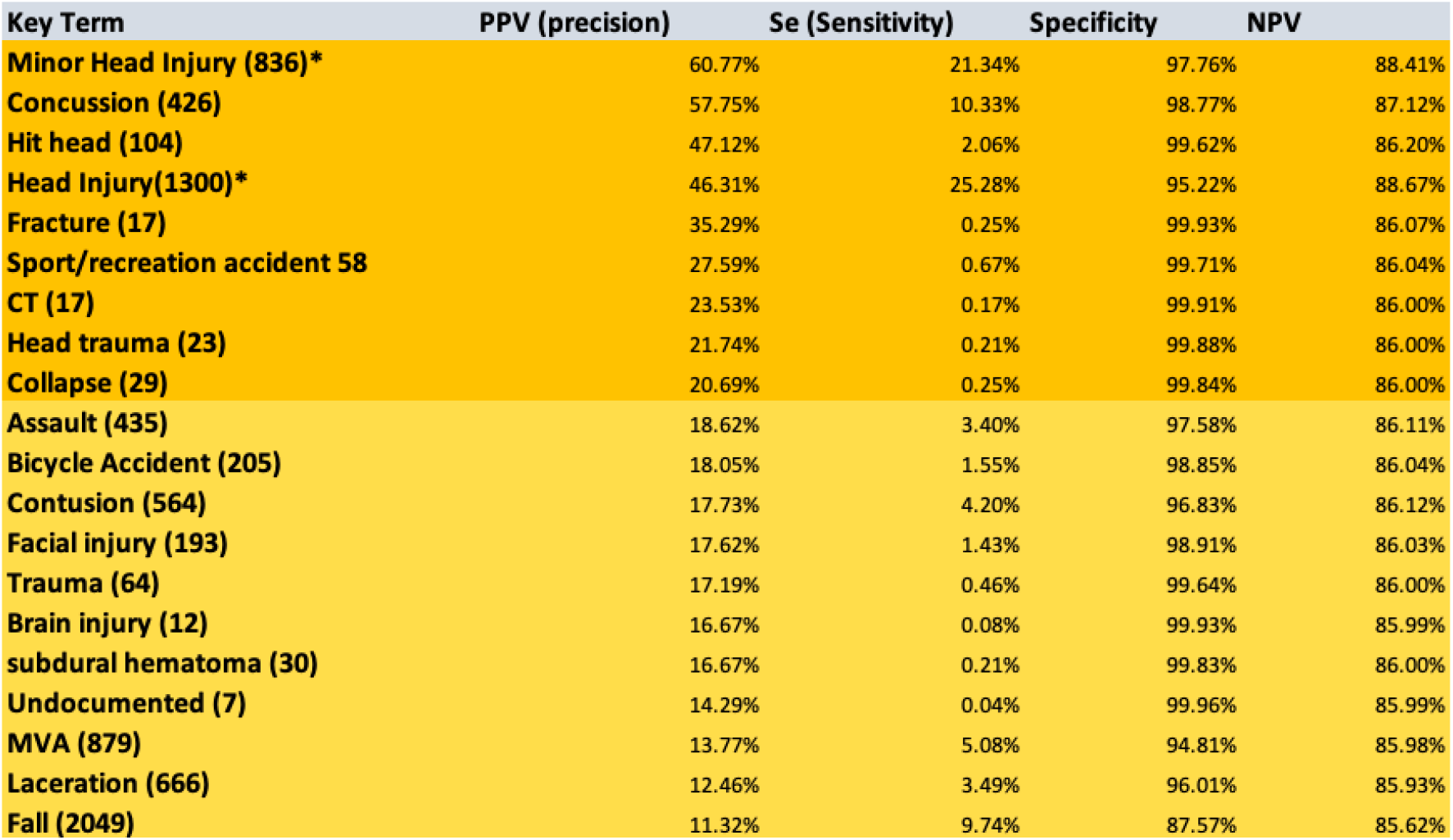
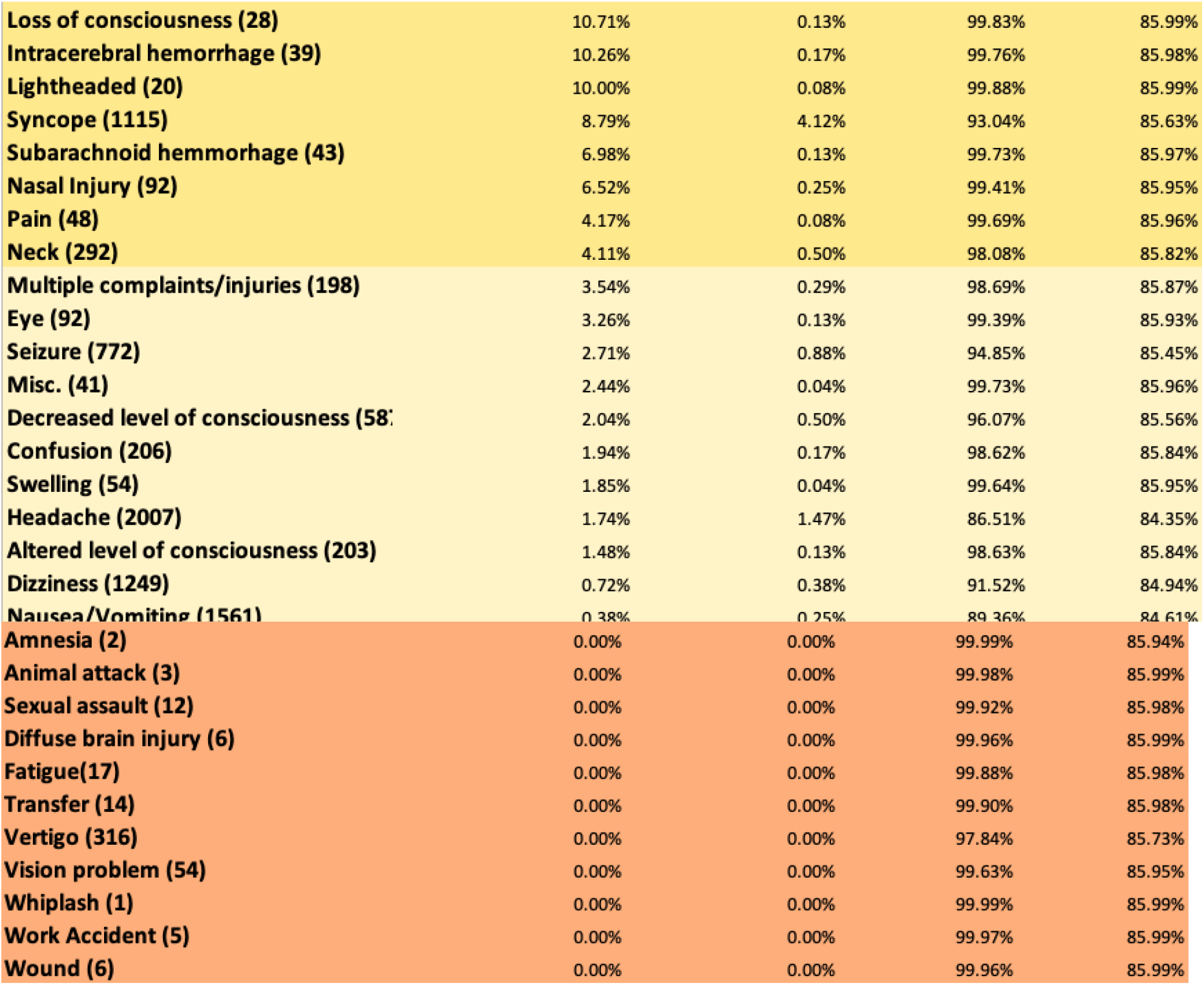
Study 1 Calculation Results for Key Terms. PPV, Se, Specificity, and NPV. Key terms are sorted by PPV. Colours indicate the five initial groupings of the key terms based on results of PPV calculations.

**Figure 5.**
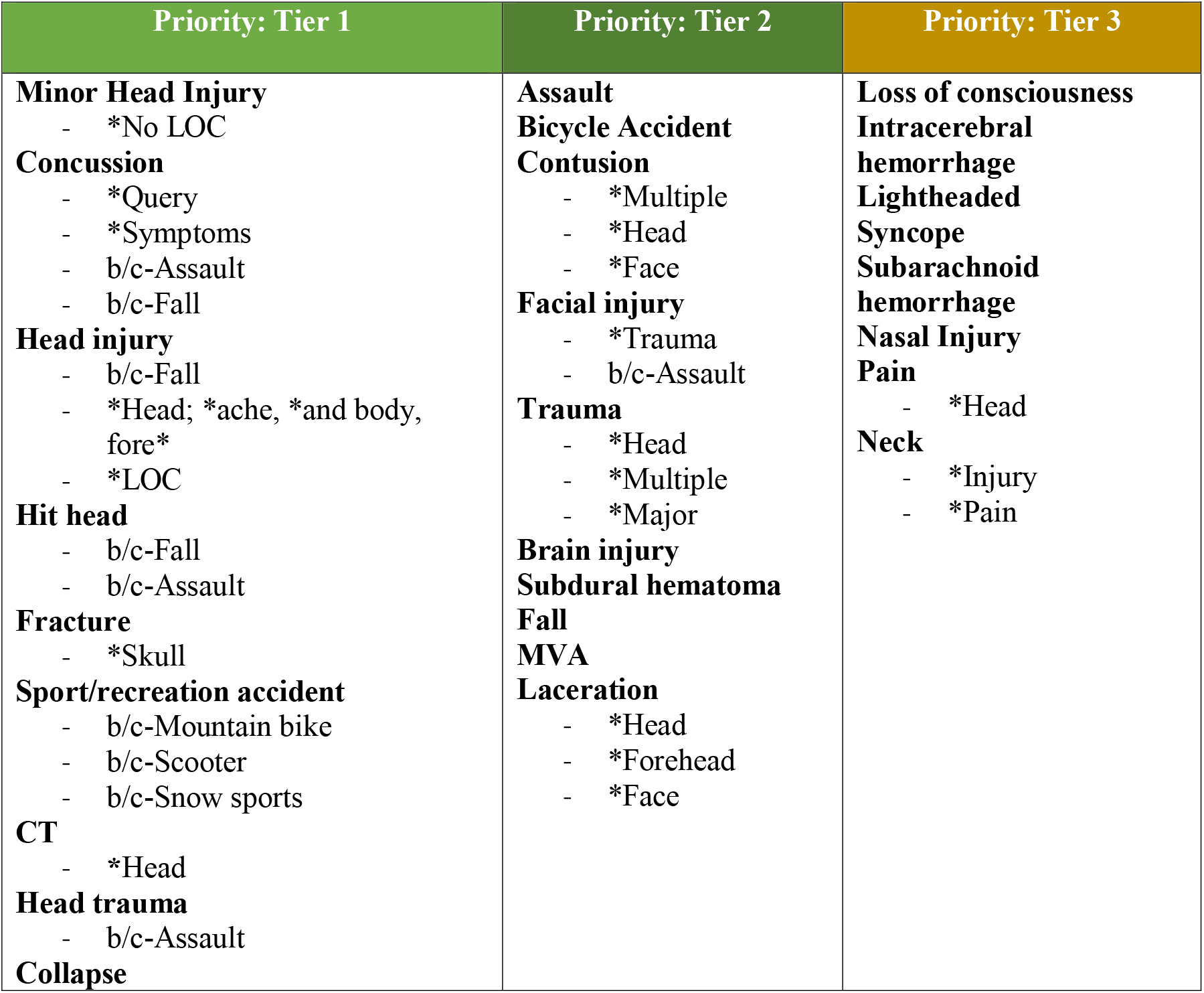
Revised Chart Review priority system. Used in study 2: “What charts do I screen?” Each column is organized from top to bottom priority. Read left to right, top to bottom. Additional terms add further specificity. Legend: * = Additional descriptor in reason for visit; b/c = bolded term because of; & = bolded key term & further descriptor in reason for visit. Chief complaints can be looked for in: CST Cerner (Powerchart): Reason for visit; PCIS: Diagnosis (ED LOG)

### Validation of the revised tiered priority chart review system

From study 2^7^, 4,434 charts were used in the sample for the analysis for the revised chart review priority system; 16 charts that were coded as a moderate/severe mTBI were excluded from the sample due to them not being relevant to this project, and 2 charts with no available chief complaint, due to RA error, were removed as well. The remaining 4,416 were used in the sample for the analysis for the revised chart review priority system and included patients’ charts who were coded as exclude, probable mTBI or indeterminate mTBI based on the chart review case ascertainment algorithm (Figure 1). We applied the same procedures for coding key terms and calculating PPV values as in study 1 (Appendix B). Note that study 2^7^ also recruited patients from outpatient clinics, but only the ED recruitment pathway was analyzed here.

Efficiency was then compared between the original priority screening system and the revised priority screening system by calculating the percent of participants found who sustained a probable or indeterminate mTBI based on the case ascertainment algorithm (Figure 1). In study 2, ED charts reviewed that did not pass the “quick screen” during chart review – meaning they either did not have a home address in BC, had no mechanism of injury, or had a pre-existing condition – were not saved into REDcap. To make the study 1 sample directly comparable, we excluded charts that did not pass the same quick screen, leaving N=4,878 charts.

In study 1, 4,878 charts that passed the quick screen were reviewed, with 2,381 people eligible to be sent an LOI. This results in a chart efficiency review rate of 48.8% being able to be sent a LOI. In study 2, 4,416 charts that passed the quick screen were reviewed, with 2,646 people eligible to be sent an LOI. This results in an efficiency rate of 59.9%. A 2-sample test for equality of proportions with continuity correction was used to compare the proportions of *people* with possible or probable mTBI based on our algorithm in study 1 vs study 2, in R. The comparison was statistically significant, χ2(1)=114.7, p<.001.

## Discussion

This study has limitations. The “true state” used to calculate PPV was the outcome of a diagnostic algorithm applied by research assistants and not a gold-standard clinical diagnosis. It is possible that some participants coded as having a possible or probable mTBI in fact did not have an mTBI, and some participants who were coded as no mTBI actually had an mTBI. This would influence the absolute PPV values but should not have impact the relative PPV differences (one key term vs. another) or the comparison between the original and revised tiered priority system. The improvement in efficiency of the revised compared to original tiered chart review priority system should be interpreted with caution because of differences between the study 1 and study 2. In study 2, charts were not reviewed from the City Center Urgent Primacy Care Center, North Vancouver Urgent Primary Care Center, and UBC Hospital. In study 2, one research assistant did all the chart review, whereas four did chart reviews for study 1. Vancouver General hospital also transitioned from PCIS (with auto-populated chief complaint fields) to Cerner (with free text fields) between study 1 and study 2, allowing us to get more specific in parsing chief complaints in study 2, and examine their PPV values more closely, which may have increased some of the PPV values of certain key terms, and decreased others.

## Conclusion

The aim of this study was to evaluate, refine, and re-evaluate a tiered priority chart review system to efficiently identifying cases of mTBI in ED/UPCCC medical charts. This iterative process yielded a system with improved efficiency – research personnel would need to review fewer medical charts to identify the same number of mTBI cases. This tiered priority chart review system could potentially be used for case ascertainment in future mTBI studies that did not require acute research data collection (e.g., blood draw for clinical purposes with hours of injury). This chart review procedure described here may be less accurate for case ascertainment than staffing the unit or clinic 24/7 with research personnel to screen patients as they arrive, but is far less resource intensive. The chart review procedure may require adaptation for settings that use different electronic medical records systems or where they may be regional differences in the terms used to characterize chief complaints.

## Data Availability

All data produced in the present study are available upon reasonable request to the authors

## Appendix A

Original tiered priority chart review system “What charts do I screen?”

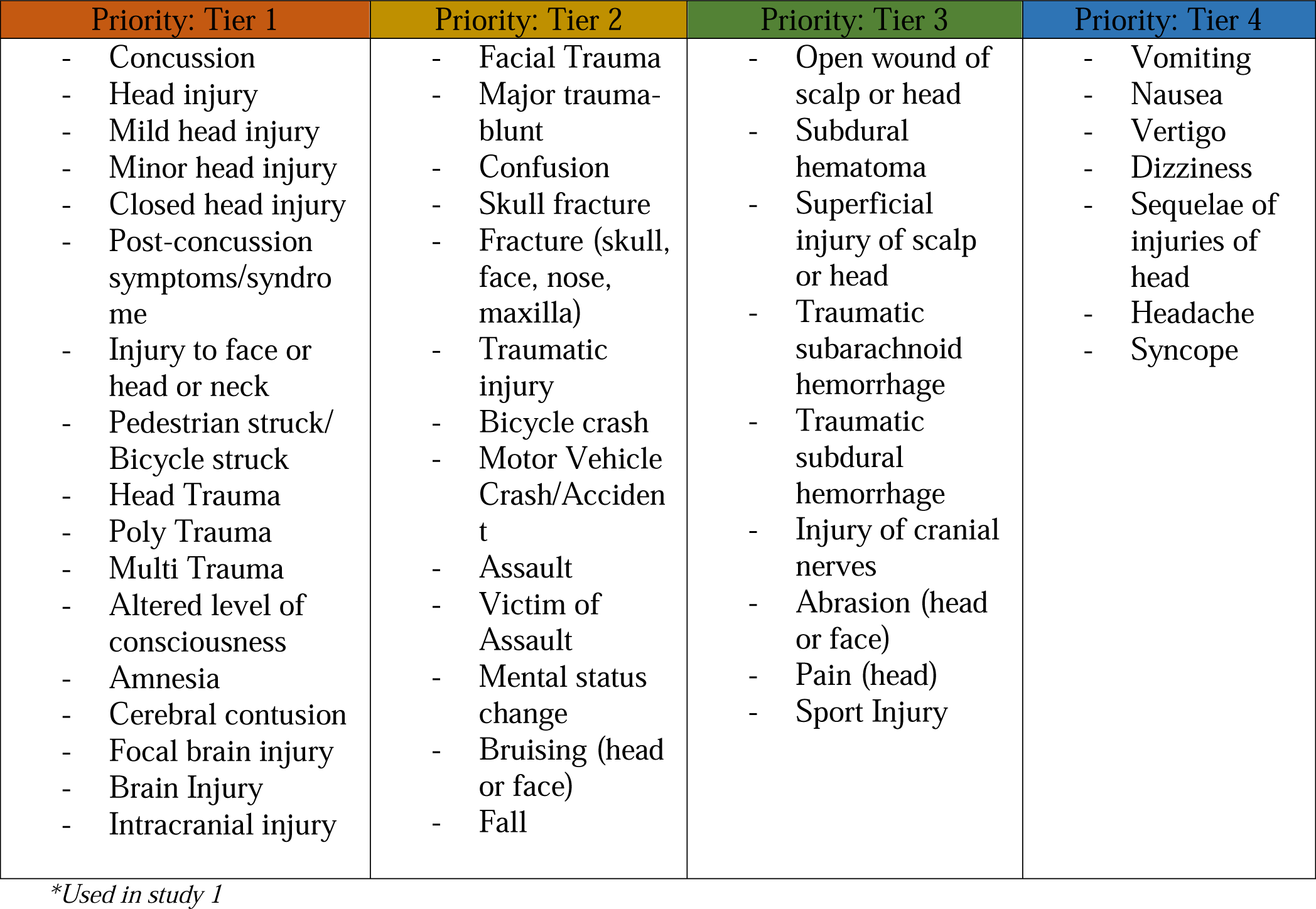

Which charts do I screen in PCIS, CST Cerner, and Intrahealth Profile?
| Priority: Tier 1 | Priority: Tier 2 | Priority: Tier 3 | Priority: Tier 4 |
| --- | --- | --- | --- |
| - Concussion<br>- Head injury<br>- Mild head injury<br>- Minor head injury<br>- Closed head injury<br>- Post-concussion symptoms/syndrome<br>- Injury to face or head or neck<br>- Pedestrian struck/ Bicycle struck<br>- Head Trauma<br>- Poly Trauma<br>- Multi Trauma<br>- Altered level of consciousness<br>- Amnesia<br>- Cerebral contusion<br>- Focal brain injury<br>- Brain Injury<br>- Intracranial injury | - Facial Trauma<br>- Major trauma-blunt<br>- Confusion<br>- Skull fracture<br>- Fracture (skull, face, nose, maxilla)<br>- Traumatic injury<br>- Bicycle crash<br>- Motor Vehicle Crash/Accident<br>- Assault<br>- Victim of Assault<br>- Mental status change<br>- Bruising (head or face)<br>- Fall | - Open wound of scalp or head<br>- Subdural hematoma<br>- Superficial injury of scalp or head<br>- Traumatic subarachnoid hemorrhage<br>- Traumatic subdural hemorrhage<br>- Injury of cranial nerves<br>- Abrasion (head or face)<br>- Pain (head)<br>- Sport Injury | - Vomiting<br>- Nausea<br>- Vertigo<br>- Dizziness<br>- Sequelae of injuries of head<br>- Headache<br>- Syncope |

*\*Used in study 1*

Which charts do I screen in PCIS, CST Cerner, and Intrahealth Profile?

## Appendix B

Study 2 Calculation Results for Key Terms PPV, Se, Specificity, and NPV of Participants Who Sustained an **mTBI**, As Found in Chart Review

|  | PPV (precision) | Se (Sensitivity) | Specificity | NPV |
| --- | --- | --- | --- | --- |
| Assault (5) | 80.00% | 0.15% | 99.94% | 39.98% |
| Minor head injury (456) | 73.03% | 12.51% | 93.05% | 41.45% |
| Head trauma (50) | 64.00% | 1.20% | 98.98% | 40.00% |
| Hit head (333) | 60.96% | 7.63% | 92.66% | 40.03% |
| Head injury (2877) | 60.88% | 76.25% | 26.37% | 42.49% |
| Loss of consciousness (5) | 60.00% | 0.11% | 99.89% | 39.96% |
| MVA (10) | 60.00% | 0.23% | 99.77% | 39.96% |
| Fall (19) | 57.89% | 0.41% | 99.55% | 39.95% |
| Concussion (590) | 57.29% | 12.70% | 85.77% | 39.54% |
| Bicycle (32) | 53.13% | 0.64% | 99.15% | 39.91% |
| Traumatic brain injury (2) | 50.00% | 0.04% | 99.94% | 39.95% |
| Sport/recreation accident (30) | 40.00% | 0.45% | 98.98% | 39.82% |
| CT (18) | 27.78% | 0.19% | 99.27% | 39.83% |
| Collapse (4) | 0.00% | 0.00% | 99.77% | 39.91% |
| Facial injury (1) | 0.00% | 0.00% | 99.94% | 39.95% |

*Study 2 Key terms. Sorted by PPV. Colours are present for increased readability, separating Key terms with PPV above 60%, between 1-60%, and below 1%.*

